# Association of LINE-1 RNA expressions in cell lines with longevity and reproductive lifespan

**DOI:** 10.64898/2025.12.17.25342501

**Authors:** Richard M. Cawthon, Ken R. Smith

**Affiliations:** Department of Human Genetics, University of Utah, Salt Lake City, UT, USA; Department of Family and Consumer Studies, University of Utah, Salt Lake City, UT, USA; Huntsman Cancer Institute, University of Utah, Salt Lake City, UT, USA

## Abstract

**Background:** Long Interspersed Nucleotide Element-1 (LINE-1, or L1) sequences occupy approximately 17% of the human genome. L1 RNA expression, required for embryogenesis, is low in middle childhood, but increases in adults, eroding heterochromatin and leading to ectopic gene misexpressions, sterile chronic inflammation, and physiological deterioration. To our knowledge, no studies have yet tested whether adults with high L1 RNA levels for their age are shorter-lived, and whether the women with higher L1 RNA levels have shorter reproductive lifespans, as would be expected if higher L1 RNA expressions accelerate both systemic and reproductive aging.

**Methods:** The RNA levels of 127 subfamilies of L1 elements in lymphoblastoid cell lines (LCLs) from 43 grandmothers and 43 grandfathers of the three-generation Utah CEPH (Centre d’Étude du Polymorphisme Humain) families were obtained from the Genetic European Variation in Disease (GEUVADIS) project. Survival and reproductive lifespan data for these subjects were obtained from the University of Utah. The sum of the RNA levels across all 127 L1 element subfamilies (a.k.a. total L1 RNA level), and the variance of RNA levels across the 127 subfamilies, were calculated for each research subject and tested for associations with longevity in both sexes and with age at last birth (ALB) for the women.

**Results:** Women in the top half of summed L1 RNA expressions, or in the top half of variance in RNA expression across the L1 subfamilies, had significantly higher mortality rates than women in the bottom half for those measures (for top half vs. bottom half total L1 RNA levels, Hazard Ratio (HR) 4.00, 95% CI 1.50-10.67, P = 0.0057; for top half vs. bottom half variance across the L1 subfamilies, HR 3.84, 95% CI 1.49-10.72, P = 0.0068). No significant associations of L1 RNA levels, or their variance, with mortality were observed in the full set of 43 men; however, restricting the analysis to the men who were 68 years or older at blood draw and survived at least four years after the blood draw (n = 31) revealed significantly higher mortality rates, within this subset of men, for those in the top half of total L1 RNA levels vs. men in the bottom half (HR 2.79, 95% CI 1.11-7.05, P = 0.03). Among the 37 women whose ALB was ≥ 30 years, the approximate age when fertility begins to decline, higher total L1 RNA levels were associated, though not significantly, with a younger ALB. However, selecting for relatively healthy individuals by restricting the analyses to women who were younger than 75.5 years at blood draw and survived at least five years after the blood draw (n = 27) revealed a strong association of higher intra-individual variance in L1 RNA expression across the 127 L1 subfamilies with a younger ALB (Pearson *r* = −0.44, *p* = 0.02).

**Conclusions:** These results from a small cohort of research subjects lend support to the hypothesis that the regulation of L1 RNA expressions in adults significantly influences the rates of both systemic and reproductive aging. Expanded studies of similar design are needed to further test this hypothesis.

## INTRODUCTION

Long Interspersed Nucleotide Elements - 1 (LINE-1 or L1) are a family of about 500,000 copies of repeated DNA elements scattered throughout the human genome that occupy approximately 17% of the genomic DNA (Wallace et al., 2008). L1 elements have been found in all mammalian species examined to date, with the exception of monotremes (Ivancevic et al., 2016). L1 RNA expression contributes critically to Zygotic Genome Activation at the beginning of embryogenesis (Li et al., 2024) at least in part by helping to open up regions of heterochromatin around thousands of genes that are silenced in mature oocytes and spermatozoa prior to fertilization. Unfortunately, L1s can also be pathogenic. About 100 L1 copies are thought to be retrotransposition-competent, and 96 cases of human genetic diseases have been reported to be due to L1 retrotransposition in the germline (Hancks et al., 2012). L1 retrotransposition also contributes to the progression of various cancers (Burns, 2017).

L1 RNA expression can be pathogenic even in the absence of retrotransposition. Several thousand distinct L1 element copies, including many that are not full-length but retain an internal promoter, undergo some level of transcription, with the frequency distribution of transcripts from different L1 subfamilies varying across tissue types and stages of development (Deininger et al., 2017), as well as between individuals (Bravo et al., 2024). L1 RNA expression is suppressed and plateaued during childhood but gradually increases with age in adults (Burattin et al., 2024), increasing more rapidly in patients with progeroid syndromes than in healthy subjects, causing erosion of heterochromatin (Della Valle et al., 2022) and consequent ectopic expression of tissue-restricted genes (Lee et al., 2021), and possibly degrading the cyclic rhythmicity of circadian gene expressions as well. Such gene misexpressions may ultimately lead to sterile chronic inflammation and physiological deterioration (Lee et al., 2021). Remarkably, delivering L1 antisense oligonucleotides (ASOs) systemically to a mouse model of Hutchinson-Gilford progeria syndrome extended the mouse’s lifespan (Della Valle et al., 2022); whether the same ASO treatment is capable of extending lifespan in otherwise normal mice has not yet been reported.

Melatonin, secreted by the pineal gland on a circadian cycle with blood levels peaking at night, suppresses LINE-1 RNA transcription *in vitro* and *ex vivo* (deHaro et al., 2014). This suggests that variation in LINE-1 RNA levels in multiple tissues may also be circadian (Belancio, 2015), possibly contributing to beneficial daily fluctuations in heterochromatin states needed to control many circadian gene expressions, including those of some housekeeping genes as well as some tissue-restricted genes; however, no studies directly testing whether L1 RNA levels in tissues fluctuate in a circadian fashion have yet been reported. Only about 5% of the body’s melatonin is produced by the pineal gland (Reiter et al., 2024). The remaining 95% is synthesized in the mitochondria of many different extrapineal cell types in response to oxidative and metabolic stresses, unassociated with circadian cycles. The suppression of L1 RNA expression by melatonin requires signaling via melatonin’s MT1 and MT2 receptors, but how much of that signaling involves melatonin of pineal vs. extrapineal origin remains unclear. Differences in L1 RNA expression after the juvenile phase of life may be an important contributor to differences in lifespan across species and to inter-individual variation in the age at death within a species.

Hypermethylation of L1 promoters strongly suppresses L1 RNA expression (Kaluscha et al., 2022). The Dog Aging Project recently found that L1 promoters are less methylated in larger vs. smaller dog breeds, suggesting that the shorter lifespans of larger dogs may be attributed, at least in part, to higher levels of L1 RNA expression (Mariner et al., 2025). The remarkably long lifespan of the naked mole rat has been attributed, at least in part, to the evolution of a mechanism for “deep silencing” of L1 elements (Kogan et al., 2024).

It is well established that physical exercise (PE) extends healthspan and lifespan (Gremeaux et al., 2012). PE also reduces L1 RNA levels in skeletal muscle (Roberson et al., 2019), increases the methylation of L1 elements in peripheral blood leukocytes (Ferrari et al., 2019), and raises blood melatonin levels (Escames et al., 2011), all of which effects are consistent with the hypothesis that the benefits of exercise are mediated, at least in part, by the lowering of L1 RNA levels.

Heritable genetic variants in humans also influence L1 RNA levels. Bravo et al. recently reported (Bravo et al., 2024) that several heritable genetic variants found in epidemiological studies to be associated with increased risks of various aging-related diseases are also associated with higher levels of L1 RNA expression in genotyped EBV-immortalized human lymphoblastoid cell lines (LCLs) from other sets of research subjects.

Those subjects included 43 grandmothers and 43 grandfathers from the three-generation Utah CEPH (Centre d’Etude Polymophisme Humain) families, whose blood samples were collected in the early 1980s to build the first comprehensive human genetic linkage map (Botstein et al., 1980; Prescott et al., 2008). Here we test whether differences in L1 RNA levels measured in this Utah cohort’s LCLs are associated with differences in those same subjects’ longevity and the female subjects’ reproductive lifespans.

### SUBJECTS, DATASETS, and METHODS

The Utah CEPH families are 47 three-generation families from Utah, each consisting of a sibship and the sibship’s parents and grandparents. These families, generally of Northern European descent, were not selected for the presence or absence of any specific disease or clinical disorder; however, the requirement for three living generations and multiple siblings in the third generation (median 8 siblings, range 4-16) may have constituted a mild selection for both longevity in the grandparents and late fertility in the middle-generation mothers.

A portion of each blood draw was used to transform B lymphocytes with Epstein Barr Virus (EBV) to generate immortalized LCLs to use as an enduring resource for the DNA needed to build the human genetic linkage map. The cell lines were deposited with the Coriell Institute for Medical Research in Camden, New Jersey so that laboratories worldwide could obtain aliquots and contribute to the mapping effort. LCLs from 82 Utah CEPH/CEU grandmothers (birth years 1892 - 1926) and 71 Utah CEPH/CEU grandfathers (birth years 1887 - 1924) are available from Coriell. Survival and reproductive history data and many additional phenotypes are available, with approval, from the CEPH/UGRP Resource for Researchers at the University of Utah (https://uofuhealth.utah.edu/center-genomic-medicine/research/ceph-resources; UGRP: Utah Genetic Reference Project, Prescott et al., 2008) for most of these 153 Utah CEPH grandparents and for many of their descendent family members as well.

EBV-induced transformation of B lymphocytes to make immortalized LCLs alters many genes’ expressions. Nevertheless, it has been shown that variation between donors in the levels of expression of thousands of genes in their LCLs is under the control of heritable genetic variants (Cheung et al., 2003; Morley et al., 2004). Furthermore, relative gene expression level differences in B lymphocytes from donors are generally preserved in their EBV-transformed lymphoblastoid cell lines (Caliskan et al., 2011). Also, normal, untransformed B-lymphocytes produce L1 RNAs (Attig et al., 2017), and lymphocytes have been reported to synthesize and secrete melatonin (Carrillo-Vico A et al., 2004). Many gene expression levels in the Utah CEPH grandparents’ LCLs are associated with the age at blood draw of the donors, and, after adjustment for age-at-blood-draw, with differences in the donors’ remaining lifespans (Kerber et al., 2009; Kerber et al., 2022); however, those studies did not include data on L1 element RNA expressions.

Here, we analyze the LCL L1 RNA levels and aging phenotypes of 43 of the Utah CEPH grandmothers (aged 57-92 years when bloods were drawn) and 43 of the grandfathers (aged 61-91 years at blood draw). L1 RNA levels from 127 subfamilies of L1 elements were collected for each LCL by the GEUVADIS (Genetic European Variation in Disease) project (Lappalainen et al., 2013) and normalized by the transcripts per million (TPM) method (Choi et al., 2025). By this method the total summed LCL L1 RNA levels across the 127 L1 subfamilies ranged from 1108 to 1363 for the women, and from 1086-1356 for the men. The VAR.S function in Microsoft Excel was used to calculate each individual’s variance in L1 RNA levels across the 127 subfamilies of L1 elements. Age at death or age last known alive for the 86 Utah CEPH grandparents, and age when the 43 women gave birth to their last live-born child, were calculated from birth dates, last-known-alive dates, and death dates obtained from the Utah Population Database (UPDB; Smith et al., 2022; Smith & Mineau, 2021), the Social Security death index, and the CEPH/UGRP Resource for Researchers (see link above). Age at death was available for 42 grandmothers (age at death: 66-101 years) and 42 grandfathers (age at death: 65-100); an age last known alive > 90 years was confirmed for each of the two remaining subjects.

We used Cox proportional hazards regression models to test whether differences in L1 RNA levels, analyzed as either a continuous or categorical variable, were associated with differences in survival. Utah had one of the highest total fertility rates in the United States throughout the first half of the 20th century. Since fertility in women declines after age 30, it is reasonable to hypothesize that among the grandmothers whose age at last birth was ≥ 30, differences in ALB can serve as a proxy for differences in their rates of reproductive aging.

## RESULTS and DISCUSSION

### Rank of subjects by L1 RNA level is preserved across L1 element subfamilies

In general, for each sex the rank of the research subjects by the RNA levels expressed from any one of the 127 L1 subfamilies was largely preserved across the various L1 subfamilies, suggesting that L1 transcription is coordinated or co-regulated genome-wide. Therefore, for our survival and reproductive lifespan analyses, we chose to examine two summary statistics for each subject: 1) their total L1 RNA level summed across all 127 L1 subfamilies, and 2) the variance of their L1 RNA levels across the 127 L1 subfamilies.

### Individual total L1 RNA level and variance of RNA levels across L1 subfamilies vs. age at blood draw

In reviewing the literature, we found no longitudinal studies examining the trajectory of L1 RNA levels within individuals during aging. However, published cross-sectional data indicate that L1 RNA levels are higher in older (>80 years old) adults than in younger (<65 years old) adults (Burattin et al., 2024, Fig. 6A). The L1 RNA data analyzed here are derived from the cell lines established from one blood draw per individual research subject, collected from subjects who were various ages at the time of their one blood draw. In these subjects neither the total RNA level in LCLs nor the variance in L1 RNA levels across the 127 L1 subfamilies varied significantly with age at blood draw in either sex. Total L1 RNA levels were ∼0.8% higher, and intra-individual variances in RNA levels across the 127 L1 subfamilies were ∼ 2% higher, in the women than in the men.

### Women with lower L1 RNA levels and lower variance in L1 RNA levels live longer

Figure 1 plots the survival function by years since blood draw for women in the top vs. bottom 50% of the distribution of total L1 RNA levels in the LCLs. The analysis is based on Cox regressions where the 43 subjects are stratified by age group (i.e. each age group has its own hazard function), and any residual mortality effects of age *within* an age group are also controlled for. The mortality rate for women in the top half for L1 RNA expression was significantly higher than for women in the bottom half (HR 4.00, 95% CI 1.50-10.67, P = 0.0057). Also, the mortality rate for women in the top half for variance in RNA expression levels across the 127 L1 element subfamilies was significantly higher than for women in the bottom half (HR 3.84, 95% CI 1.49-10.72, P = 0.0068).

**Figure 1.**
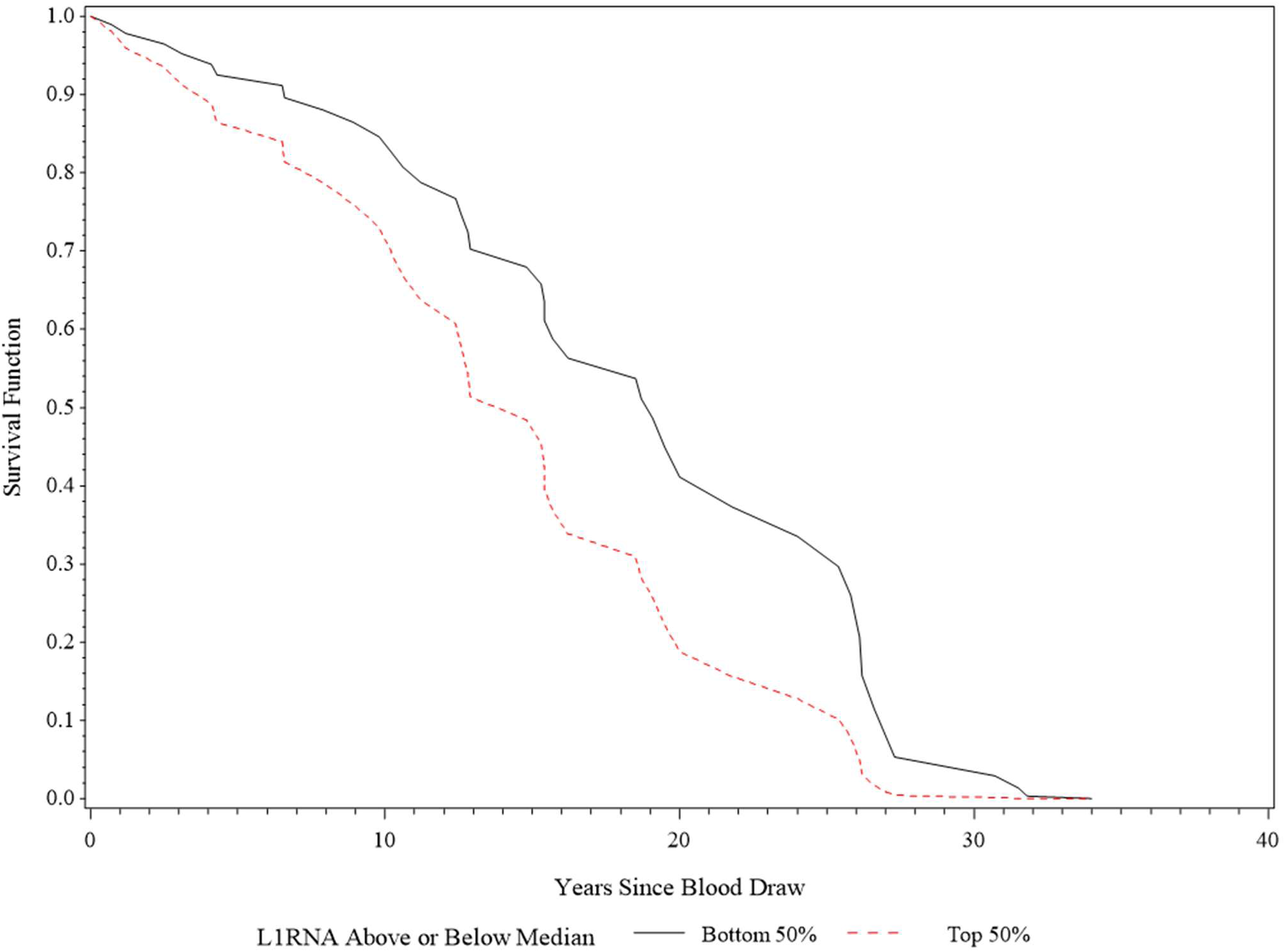
Survival of 43 women by total L1 RNA expression levels. The difference in median survival for the two groups was approximately 5.2 years.

### Men with lower L1 RNA levels live longer

In the full set of 43 Utah CEPH grandfathers, we found that higher total L1 RNA levels were associated with shorter survival, though not significantly. Reasoning that the 5 men who survived less than 4 years after the blood draw may have had a terminal illness affecting L1 RNA levels, we excluded those men from subsequent analysis. Note that physical exercise (PE) increases methylation of L1 elements in peripheral blood leukocytes (Ferrari et al., 2019), likely suppressing L1 RNA expression (Kaluscha et al. 2022). Since PE declines steeply in men in their 60s (Martin et al., 2014; Jefferis et al., 2015), we reasoned that stable LCL L1 RNA differences between men, potentially associated with differences in their mortality risks late in life, may be more easily detected in men whose bloods were drawn at ages > 68 years, when the men are more likely to be sedentary. After applying these two sets of exclusion criteria, 31 men remained for analysis of survival vs. L1 RNA levels.

Figure 2 plots the years since blood draw vs. survival for the men in the top vs. bottom 50% of the distribution of total L1 RNA levels in the LCLs. The analysis is based on Cox regressions where age was adjusted using age at draw and age at draw squared (i.e., allows for non-linear effects of age on mortality). Age stratification, as done with the female sample, proved unstable given the smaller male sample. The mortality rate for men in the top half for L1 RNA expression was significantly higher than for men in the bottom half (HR 2.79, 95% CI 1.11-7.05, P = 0.03). Here, age is controlled for at its mean (∼75 years).

**Figure 2.**
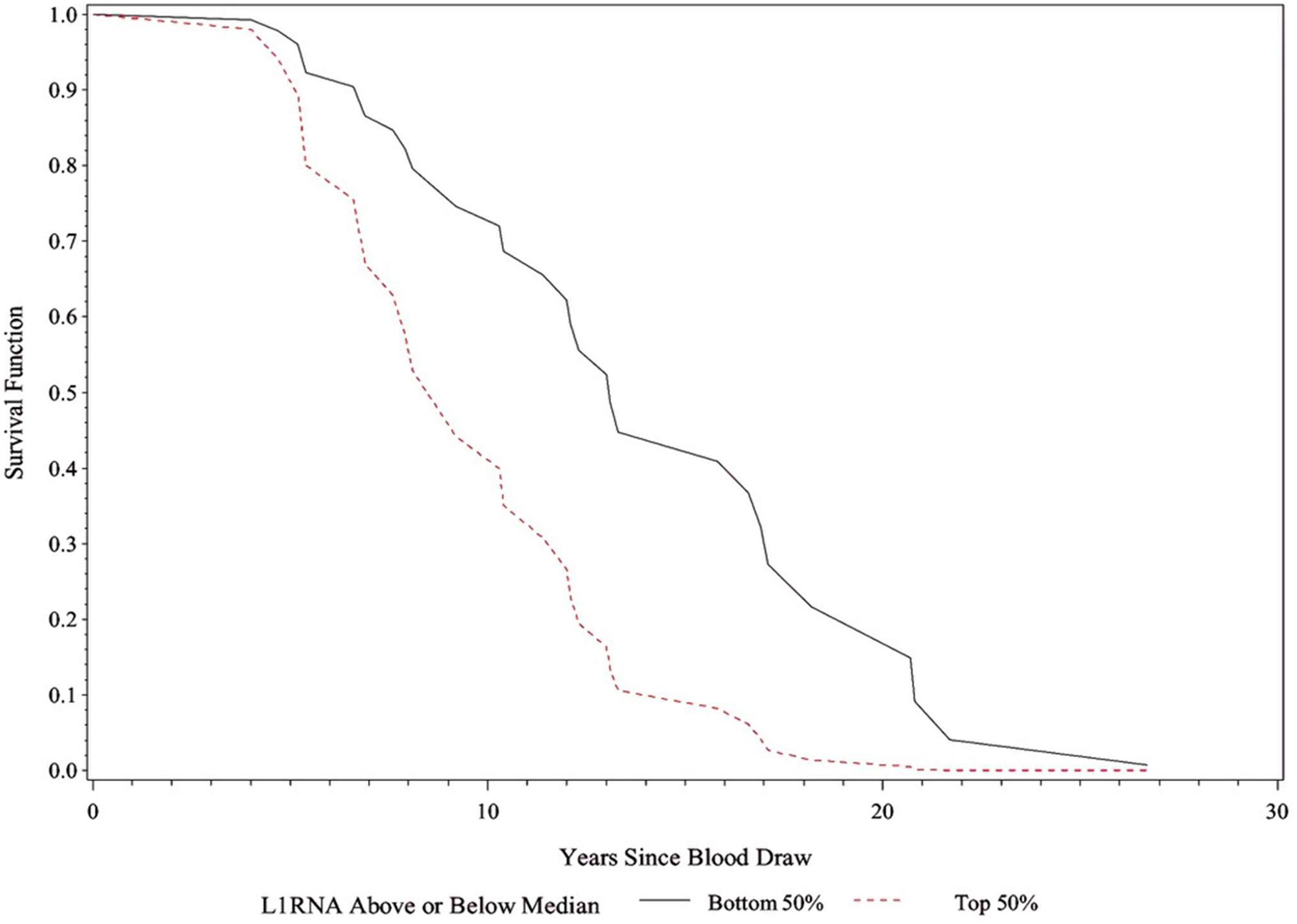
Survival of 31 men by total L1 RNA expression levels. The difference in median survival for the two groups was approximately 4.6 years.

### Women with lower variance in L1 RNA levels maintain fertility longer

Since fertility in women does not begin its decline until approximately age 30, variation in age at last birth (ALB) before age 30 is unlikely to reflect differences between women in the rate of reproductive aging. Therefore, we excluded from the L1 RNA vs. ALB analysis the six women with ALB < 30. For the remaining 37 women with ALB ≥ 30, higher L1 RNA levels were indeed associated with younger ALBs, as our hypothesis predicted, but the association did not reach statistical significance. We then applied additional exclusion criteria with the aim of removing subjects who at the time of their blood draw may have had a terminal illness or late-onset frailty condition (e.g. chronic disability and/or disease) that could potentially influence the L1 RNA levels in their blood cells, thereby possibly confounding our analyses. Frailty in women increases significantly after approximately age 75 (Lee et al., 2018, Fig. 2). Therefore, we excluded subjects age ≥ 75.5 years at blood draw and subjects who died less than 5 years after the blood draw. After applying these additional exclusions, 27 women remained for the L1 RNA levels vs. ALB analysis.

Figure 3 Panel A shows that a higher total L1 RNA level was associated, though not significantly, with a younger ALB. In Panel B, we see that intra-individual variance in L1 RNA levels across the 127 L1 subfamilies was significantly associated with ALB. Panel C shows that the ratio: (Variance across the L1 subfamilies) / (Total L1 RNA) was more strongly associated with ALB than either the total L1 RNA level (Panel A) or the variance in L1 RNA levels across L1 subfamilies (Panel B). These results suggest that lower total L1 RNA levels and lower variance in RNA expressions across the various L1 elements both contribute importantly to the maintenance of fertility in women as they age.

**Figure 3.**
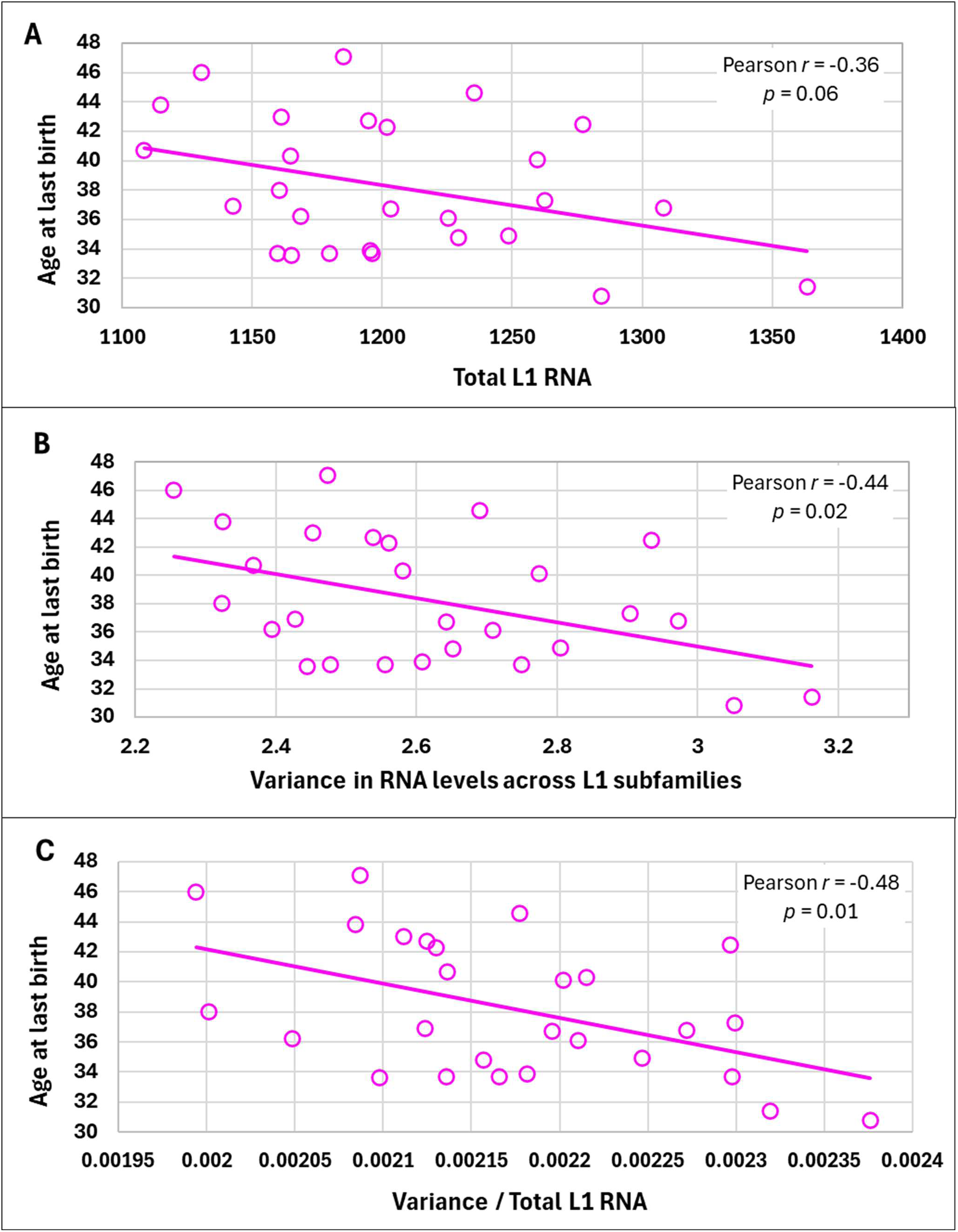
L1 RNA expression vs. reproductive lifespan of 27 women. Here, age at last birth (y axes) is plotted against three different LCL L1 RNA parameters (x axes): **A,** total L1 RNA levels; **B,** variance in the RNA expression across 127 L1 element subfamilies; and **C,** the ratio of that variance to total L1 RNA.

## CONCLUSIONS

Here we report that L1 RNA transcription levels in cultured LCLs from middle-aged or older donors are associated with survival in both sexes and the duration of fertility in women. Thus, the effectiveness with which L1 RNA levels are controlled may be an important determinant of the rates of both systemic and reproductive aging. We view these observations, analyses, and interpretations as preliminary and exploratory. Similar studies of additional, larger populations are needed to test the reproducibility of our results.

Melatonin suppresses L1 RNA expression (Belancio, 2015). Plasma melatonin concentrations during the daytime are very low across the lifespan. In contrast, plasma melatonin concentrations during the night are highest during childhood, fall dramatically during puberty and adolescence, and decline further in adults, particularly after age 60 (Kennaway, 2023). Nighttime plasma melatonin levels in young women are higher than in young men; women’s melatonin levels then decline before, during, and after menopause, when they reach levels below those of similarly aged men (Gursoy et al., 2015; Obayashi et al., 2015). Therefore, it is plausible that while melatonin’s circadian nighttime suppressions of L1 RNA during childhood, in combination with other defenses, may maintain childhood’s robust homeostatic systems, minimizing the accumulation of damage to the genome and other cellular components, the hormone’s decline at puberty and beyond would be expected to erode circadian rhythms and allow increasingly sustained L1 RNA expressions to cause genomic and cellular damage to accumulate, ultimately resulting in the decline in fertility in women with age and the progression to frailty and mortality in both sexes with which we are all familiar.

Consistent with the above perspective, melatonin given to mice extends the lifespan of both female and male mice (Pierpaoli et al., 1991; Damiani et al., 2020) and delays ovarian aging in female mice (Tamura H et al., 2017). Melatonin supplements reduce the levels of chronic inflammation in adults (Cho et al., 2021). Several clinical trials are underway to determine whether melatonin supplementation in adults can delay and/or ameliorate multiple aging-related frailties and diseases. Melatonin supplements may slow aging in at least three ways: by suppressing L1 RNA expression (Belancio, 2015), lowering body temperature (Marrin et al., 2013), and improving circadian rhythmicity (Cajochen et al., 2003). It remains to be seen whether regular melatonin use extends healthspan and lifespan in adults, and if it does, how much of those effects will be attributable to the suppression of L1 RNA expression.

### Future Directions

Since LCLs clearly retain gene expression profiles that are informative for the rates of aging of the blood donors, it is reasonable to screen libraries of already FDA-approved compounds for their ability to shift the gene expression profiles of LCLs from fast-aging subjects towards the profiles found in LCLs from slow-aging subjects. Several tests that measure the rate of biological aging from a blood sample are available (see, for example, Belsky et al., 2020). The discussions above suggest that melatonin, alone or in combination with other compounds, would be a top candidate compound to be screened in this way. Because many compounds that promote healthy longevity may operate via signaling pathways that naturally operate on a circadian rhythm, compounds with a short half-life in the body would be the preferred screening set; once a compound or combination of compounds advances to pre-clinical trials in mice and dogs, with healthspan and lifespan as the outcomes of interest, the trials should include arms testing the possible influence of time of day of administration on safety and efficacy.

## Data Availability

All data produced in the present study are available upon reasonable request to the authors and approval by the University of Utah Institutional Review Board, Resource for Genetic and Epidemiologic Research, and Technology Licensing Office.

Life history and clinical outcomes data. Utah Population Database (UPDB) data contributed to this project using birth and death records and family data that include Protected Health Information and individual identifiers. Special attention is given to protect individuals and their information contained within the UPDB and the organizations that contribute data while also allowing access to researchers. Accordingly, the Utah Resource for Genetic and Epidemiologic Research (RGE), established in 1982 by Executive Order of the Governor of Utah, administers access to the UPDB through a review process of all proposals using UPDB data. The protection of privacy and confidentiality of individuals represented in these records has been negotiated with agreements between RGE and data contributors. Data from the UPDB is available only for approved health-related research studies and access is project-specific and granted after review and approval by an RGE oversight committee and the University of Utah’s IRB. This process allows researchers with approved protocols to use the data, a process that has proven effective and successful as evidenced by hundreds of approved studies that have relied on the UPDB. Editors and reviewers of our manuscript who feel that they need access to UPDB source data beyond that which is contained in the manuscript, in order to evaluate our study, are requested to contact the corresponding author. Once those needing access sign an RGE Confidentiality Agreement and the request for data access is promptly reviewed and approved, access to the relevant data will be granted.

## ACKNOWLEDGEMENTS

We thank all the Utah individuals who participated in the CEPH consortium and all family members who participated in the UGRP. We thank Ray White, Ph.D. (deceased), and Jean-Marc Lalouel D.Sc., for their leadership in ascertaining and enrolling the Utah CEPH families in the 1980s to build the first comprehensive human genetic linkage map; and Stephen M. Prescott, M.D. (deceased) for his leadership in envisioning and building the UGRP. We thank the Pedigree and Population Resource of the Huntsman Cancer Institute, University of Utah (funded in part by the Huntsman Cancer Foundation) for its role in the ongoing records collection, maintenance and support of the UPDB. We thank the UPDB and UGRP staffs and Huong Meeks, Ph.D. for performing the updates and verification of Utah CEPH survival and reproductive histories data that we used for a previous study (Cawthon et al., 2020) and again in the present study. We thank Richard A. Kerber and Elizabeth O’Brien for helpful discussions and suggestions. We thank Bérénice A. Benayoun Ph.D. and Juan I. Bravo, Ph.D., University of Southern California, for helping us to access the GEUVADIS LINE-1 RNA expression levels for the Utah CEPH family members.

## FUNDING

This work was supported by NIH P30CA2014 (to the Utah Population Database, a.k.a. the UPDB); National Center for Research Resources Public Health Services grant M01RR00064 (to the Huntsman General Clinical Research Center, University of Utah); National Center for Advancing Translational Sciences NIH grant UL1TR002538 (to the University of Utah’s Center for Clinical and Translational Science); gifts from the George S. and Delores Doré Eccles Foundation (to the University of Utah) that supported the Utah Genetic Reference Project (UGRP); and NIH grant R01AG022095 (to K.R.S.).

## AUTHOR CONTRIBUTIONS

R.M.C. conceived the idea for the study and wrote the original draft. K.R.S. and R.M.C. performed all statistical tests of the association of the lymphoblastoid cell lines’ LINE-1 RNA expression levels, obtained from the GEUVADIS project, with the aging-related phenotypes provided by the UPDB and the UGRP. Both authors provided critical feedback and helped to shape the research and data analysis, and the review and editing of the manuscript.

## COMPETING INTERESTS

The authors declare that they have no potential conflicts of interest relevant to this study.

## ETHICS STATEMENT

All participants provided signed, paper-based informed consent. The University of Utah’s Institutional Review Board (IRB) and the University of Utah’s Resource for Genetic and Epidemiologic Research (RGE) have approved this study for publication (IRB_00021454, IRB_00011975, and RGE_00000020).

## Notes

### Competing Interest Statement

The authors have declared no competing interest.

### Funding Statement

This study was funded by NIH P30CA2014, M01RR00064, UL1TR002538, gifts from the George S. and Delores Dore Eccles Foundation, and NIH grant R01AG022095 (to KRS).

### Author Declarations

The University of Utah's Institutional Review Board has reviewed and approved our study, "Association of LINE-1 RNA expressions in cell lines with longevity and reproductive lifespan", in its entirety.

